# Next-generation sequencing of pancreatic cyst wall specimens obtained using Moray micro-forceps for improving diagnostic accuracy

**DOI:** 10.1101/2023.02.23.23286230

**Authors:** Stuart Astbury, Aishwarya Baskar, Jane I Grove, Philip Kaye, Aloysious D Aravinthan, Martin W James, Christopher Clarke, Guruprasad P Aithal, Suresh Vasan Venkatachalapathy

## Abstract

**Background and study aims:** Pancreatic cysts are common incidental findings, with an estimated prevalence of 13-15% in imaging done for other reasons. It is difficult to identify cysts with malignant potential. Diagnosis often relies on collection of cyst fluid, but tissue sampling using micro-forceps may allow for a more reliable diagnosis and higher yield of DNA for next-generation sequencing (NGS).

**Patients and methods:** 24 patients referred for endoscopic ultrasound were recruited. Biopsies were taken using micro-forceps and the AmpliSeq Cancer Hotspot panel was used for NGS, a PCR assay targeting several hotspots within 50 genes, including GNAS, KRAS and VHL.

**Results:** The concentration of DNA extracted from 24 cyst wall samples was significantly higher than in the 9/24 available matched cyst fluid samples. Cyst wall biopsy was able to diagnose 19/24 cysts (5 high risk, 6 intraductal papillary mucinous neoplasm and 4 benign). The sensitivity, specificity and diagnostic accuracy for standard of care was 66.6%, 50% and 63.1% respectively and for standard of care with NGS was 100%, 50% and 89.4% respectively.

**Conclusions:** Cyst wall biopsy performs well in diagnosing cysts but was inadequate in 5/24 patients. NGS data correlates well with histology and may aid in diagnosis and risk stratification of pancreatic cysts.

## Introduction

Pancreatic cancer is the fourth leading cause of death with a median five-year survival of 4% [1], the overall median survival is 4.6 months from diagnosis [2]. Early diagnosis of pancreatic cancer remains a major challenge and is one of the root causes of the lack of improvement in outcomes of pancreatic cancer worldwide [3]. The Cancer of the Pancreas Screening Study (CAPS3) revealed that 10% of the population screened with a family history of pancreatic cancer had intraductal papillary mucinous neoplasms (IPMN) [4]. In a secondary screening programme involving the European Registry of Hereditary Pancreatitis and Familial Pancreatic Cancer (EUROPAC), cystic lesions were the most common finding of which more than half were IPMNs, although these were independent of genetic predisposition [5]. In this group of patients, earlier diagnosis of high-risk cysts may change their treatment and therefore outcome.

The incidence of pancreatic cysts is 2.5% in the general population but steadily increases to 10% in patients over the age of 70 [6, 7]. Approximately 15% of patients who undergo computed tomography (CT) or magnetic resonance imaging (MRI) for unrelated reasons will have a pancreatic cyst [8]. Common pancreatic cysts include serous cystadenoma (SCA), pancreatic pseudocyst, mucinous cyst neoplasm (MCN) and intraductal mucinous papillary neoplasm (IPMN). SCA and pancreatic pseudocysts are non-mucinous and do not carry malignant potential. However, MCN and IPMN have the potential for malignant transformation (35-50%) and the annual malignant transformation risk is 2%, necessitating regular follow-up [9, 10].

At present, there is no single investigation that will accurately differentiate between high-risk cysts that need intervention, intermediate cysts that require surveillance and low risk cysts, which do not require further surveillance. The current standard of care for patients with pancreatic cysts is to have either CT or MRI with or without endoscopic ultrasound (EUS) and cyst fluid assessment for carcinoembryonic antigen (CEA), amylase and cytology when a sufficient amount of cyst fluid can be aspirated [11].

Next-generation sequencing (NGS) of cyst fluid has shown promise in improving the diagnostic accuracy of differentiating pancreatic cysts [12]. Singhi et al. extracted DNA from cyst fluid and used a deep sequencing panel, targeting mutations in KRAS/GNAS and TP52/PIK3CA/PTEN to identify IPMN and advanced neoplasia, respectively. Other studies have demonstrated the utility of NGS for stratifying pancreatic cysts, potentially biasing results towards cysts that more readily shed DNA [13-15]. Additionally, aspiration of adequate fluid through fine needle aspiration (FNA) needle for cytology and DNA extraction is more challenging as the viscosity of cyst fluid increases [16].

The Moray micro-forceps are biopsy forceps that can be passed through a 19-gauge fine needle aspiration (FNA) needle, allowing for sampling of pancreatic cyst wall tissue [17]. We conducted a prospective observational study to test the effectiveness of the Moray micro-forceps at increasing the amount of DNA available for NGS relative to that of cyst fluid, and whether this could provide an aid to standard histological and radiological diagnosis. We evaluated the use of cyst wall biopsies obtained with Moray micro-forceps compared to cyst fluid sampling for earlier detection of somatic variants indicating potential for cancer development, and whether this could aid in the diagnosis of progressive cyst types.

## Materials and methods

This was a single centre, prospective, observational cohort study. Patients were recruited from Nottingham University Hospitals NHS Trust. Ethical approval was obtained from the Nottingham Health Science Biobank access committee (Reference: 15/NW/0685, approval ACP000282). All adult patients with pancreatic cyst(s) ≥ 1.5 cm diameter who were referred for EUS assessment from the regional hepato-pancreato-biliary multi-disciplinary meeting between 15 January 2019 and 15 June 2020 were eligible for inclusion. Patients with a cyst size of less than 1.5cm, or who were clinically diagnosed with pseudocysts were excluded from the study. Patient demographics, cytology, histopathology, imaging findings, CEA analysis, fluid amylase, and EUS findings were documented.

### Primary and secondary objectives

The primary objective of the study was to identify whether a larger quantity of DNA suitable and sufficient for NGS can be obtained from the cyst wall compared to cyst fluid. The secondary objectives were to assess (1) if NGS alone or in combination with standard of care (CT/MRI + EUS assessment ± fluid CEA ± fluid amylase) improves the sensitivity, specificity and diagnostic accuracy of cyst identification, (2) adequacy of samples for histological assessment, and (3) the complications associated with the cyst wall biopsy using Moray micro-forceps.

### EUS procedure and cyst wall biopsy

The procedure was carried out under conscious sedation using a combination of fentanyl and midazolam. EUS was performed by independent endo-sonographers who had received formal training in the fine needle aspiration (FNA) technique. Using EUS, the cyst was identified, evaluated for any high-risk features (>3cm, dilated pancreatic duct >9mm, enhancing wall, mural nodule) and the most accessible portion of the cyst adjacent to the mucosa on EUS was selected. A 19-gauge FNA needle was then introduced into the cyst with 5-10ml dry suction. Fluid was aspirated where possible, allowing the cyst to partially collapse and was sent for amylase, carcinoembryonic antigen (CEA) and the remainder snap-frozen in liquid nitrogen for DNA extraction. Through the FNA needle, Moray micro forceps were introduced into the cyst, opened under ultrasound guidance, withdrawn close to the needle tip, needle tip advanced to the opposite wall, biopsy forceps closed, needle gently withdrawn until there was denting of the cyst wall and then the biopsy forceps were removed in a single motion. This was repeated 3-5 times and intravenous antibiotics were given at the end of the procedure. Samples were placed in formalin for routine histology and excess tissue was immediately snap-frozen in liquid nitrogen and stored at -80°C.

### Histopathological and radiological analysis

The CT/MRI images were reported by an expert independent radiologist blinded to clinical information, previous radiology reports and laboratory results. Similarly, histology was reported by an independent histopathologist blinded to the clinical, radiological and laboratory biochemical information. The slides were prepared at Nottingham University Hospitals NHS Trust, stained for haematoxylin & eosin and sent to the histopathologist with an anonymised numeric code.

### DNA extraction and NGS of cyst wall biopsies and cyst fluid

DNA was extracted from cyst wall samples using the Qiagen QIAmp DNA Micro kit and quantified using a Qubit Fluorometer. PCR amplicons were generated using the AmpliSeq for Illumina Cancer Hotspot Panel v2 (primer sequences in Supplementary table 1) and sequenced on an Illumina MiSeq. Demultiplexed paired end reads were filtered for erroneously short and long sequences, low quality scores (<Phred 30) and adapter sequences using Trimmomatic. Alignment to the human genome reference hg38 was carried out using bwa-mem [18]. The remaining processing was carried out following GATK best practices [19], alignments were filtered using BaseRecalibrator, and somatic variants were called using Mutect2 and FilterMutectCalls [20]. As healthy cyst-adjacent pancreatic tissue from each participant was not available as a control, a panel of normal samples was constructed using publicly available data from the 1000 Genomes project [21] to filter alignment artifacts, and the gnomAD database [22] was used to filter known germline variants. Finally, the filtered list of variants for each sample was annotated using SNPeff [23] to identify coding and non-coding variants. The limit of detection used was a mutant allele frequency (MAF) of 1%, with a minimum sequencing depth of 7000x. Due to the small sample size the list of targets used for analysis was limited to genes with hotspots previously linked to pancreatic malignancies according to the Catalogue of Somatic Mutations in Cancer (COSMIC) database [24].

Confirmatory NGS was carried out on cyst fluid where samples were available. DNA was extracted using the Qiagen ccfDNA extraction kit and primers from the AmpliSeq panel were used to amplify relevant amplicons (these sequences are highlighted in supplementary table 1). Amplified DNA was cleaned using the Qiagen PCR cleanup kit and normalised to 20ng/µl. Samples were sequenced by Azenta/GENEWIZ (Essex, UK). Raw sequence data was processed using an identical workflow to that described above for the cyst wall samples.

### Case definition

Four patients in the study had surgical resection. The cyst wall biopsy specimens were compared against the resected surgical specimen. The cyst wall biopsy correlated well with the surgical specimen. Hence, cyst wall tissue histopathology was used as the standard and all parameters were compared against histopathological diagnosis. Standard of care (SOC) is defined as patients having CT/MRI ± EUS± Cytology ± Fluid CEA/amylase to identify if they have high-risk cysts.

### Statistical analysis

Categorical variables were presented as numbers with percentages; continuous variables were presented as median and Interquartile range (IQR). Statistical analyses were performed using Prism 9 (GraphPad Inc., San Diego, CA) and SPSS Statistics for Windows, Version 28.0 (IBM, Armonk, NY). A p value <0.05 was considered to indicate statistical significance. Diagnostic accuracy for individual diagnostic test/parameters was calculated using sensitivity, specificity, positive predictive value, negative predictive value, positive likelihood ratio, and negative likelihood ratio.

## Results

### Clinical characterisation of patients

Twenty-four patients (54% female) were prospectively recruited to the study (Table 1); the median age was 71 years (IQR 59 – 78). Sixteen patients (67%) were of European descent, 1 (4%) was Asian and for 7 (32%) the ethnicity was not known. After the procedure, patients were followed up for a median of 491 days (IQR 126 – 674).

**Table 1:**
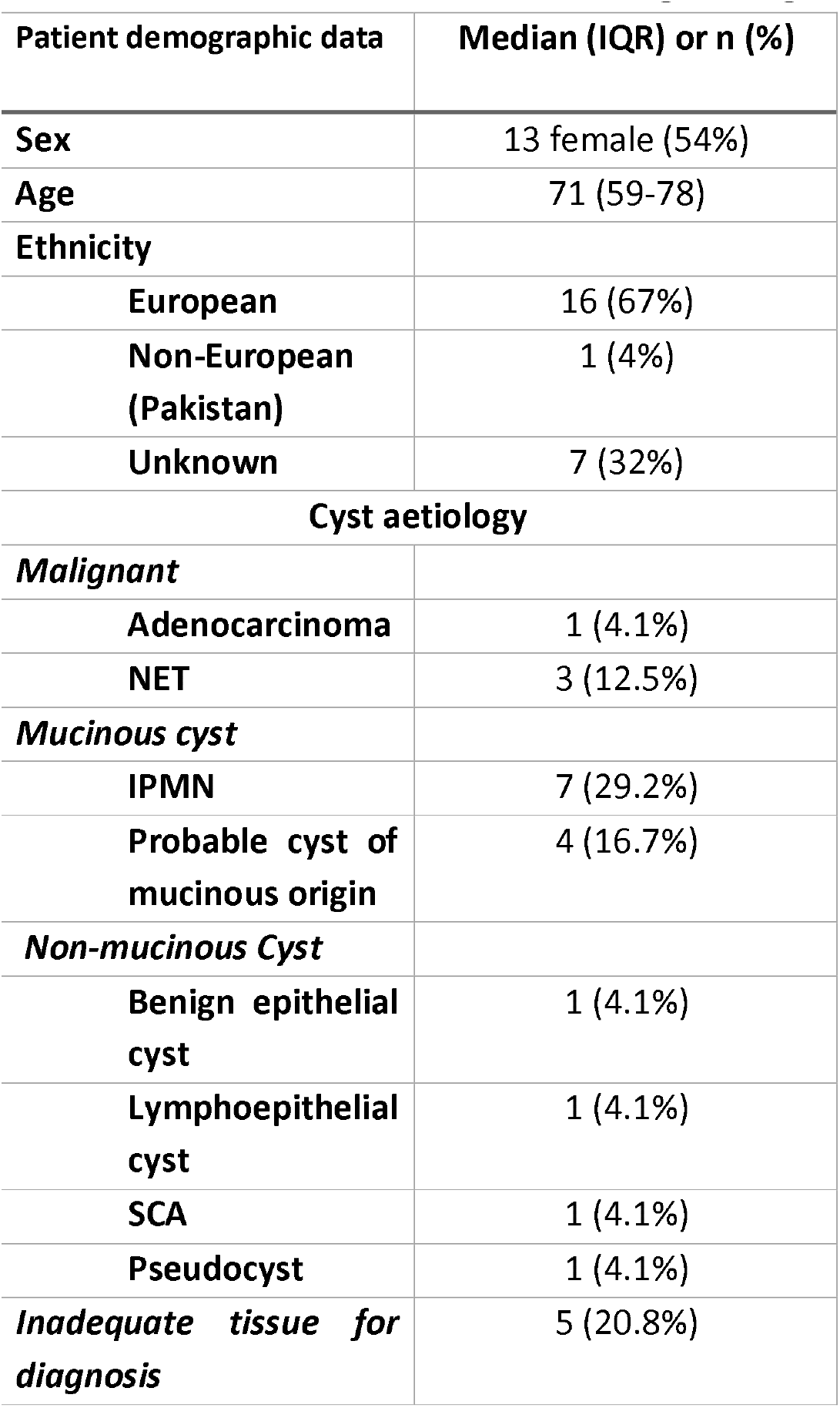
Cohort characteristics and histological diagnosis based on cyst wall biopsy.

### Histological analysis

The histopathologist was able to give a definite diagnosis based on H&E staining in 15/24 (62.5%) cases, probable diagnosis in 4/24 (16.6%) cases, and in 5/24 (20.8%) of cases the sample was inadequate for diagnosis. The histological outcomes are shown in Table 1. Cyst wall biopsy specimens were compared against resected surgical specimens in four available cases.

#### Radiological analysis

22/24 (91.7%) patients had either a CT or MRI scan. In 2 patients the CT/ MRI was performed in referring hospitals and the images could not be retrieved for independent evaluation. Communication with the pancreatic duct (PD) was established only in 6 patients (27.7%). The main PD was visualised in 14/22 patients (63.6%) and the median diameter was 3mm (IQR 1-2). Mural nodules and septations were identified in 4 (18.1%) and 3 (13.6%) patients, respectively. 15/22 (68.2%) were reported as side branch IPMN (SB-IPMN), 5 as MCN, 1 SCN and 1 adenocarcinoma.

### EUS and fluid cytology

The median diameter of the PC was 25mm (IQR 16 – 33). The median PD diameter was 2mm (IQR 2 – 3.5) and communication of the cyst with PD was defined in 23 patients (95.8%). Mural nodules and septations were identified in 4/24 (16.6%) and 7/24 (29.1%), respectively. Obtaining fluid for cytology from the cyst was feasible in 16/24 (66.6%) of patients, but definite diagnosis from fluid cytology was possible in 4/16 (25%) of these patients. The remainder had either bland epithelium or the sample was acellular. Aspirated cyst fluid was sent for CEA and amylase analysis. CEA analysis was possible in 7/24 (29.1%) and amylase in 12/24 (50%) of patients.

### Follow-up and serious adverse events

One patient developed acute severe pancreatitis requiring prolonged hospitalisation. There were no other complications secondary to the procedure and there were no deaths within 30-days of the procedure. Follow up information was available for all patients. The median follow up period was 491 days (IQR 125 – 674). Four patients had surgical resection, 2 patients had successful radiofrequency ablation of the cyst, 1 had progression of the cyst, 7 had stable cysts, 4 were discharged with benign cysts, 3 were discharged after intervention and 3 died (2 within 12 months and 1 within 2 years). The patients who died had adenocarcinoma (126 days-progression of cancer), IPMN with low-grade dysplasia (74-days-Sepsis) and IPMN with high grade dysplasia (514 days-Progression to cancer).

### Evaluation of DNA yield from samples

Of the 16 cyst fluid samples collected, sufficient DNA for NGS was obtained from 9 samples; whereas sufficient DNA was extracted from 24/24 (100%) of cyst wall biopsy samples. The median concentration of DNA from cyst fluid and cyst wall was 1.03 ng/µl (IQR 0.28 – 5.79) and 7.71 ng/µl (IQR 4.74 - 27.5), respectively (Figure 2). The difference between the DNA quantity obtained from the cyst wall tissue and the cyst fluid was significant (p=0.003, Mann-Whitney U test).

**Figure 1:**
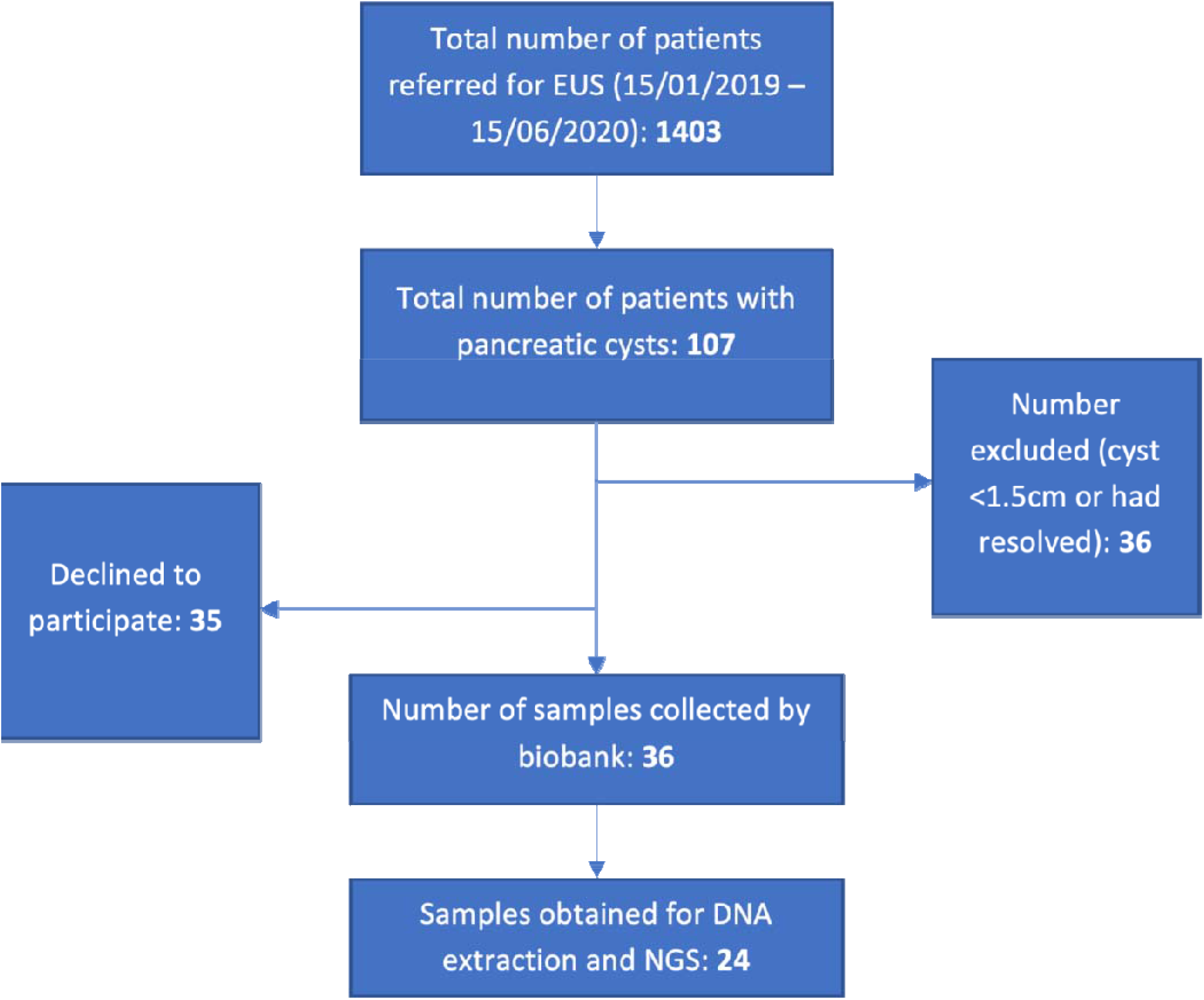
Study flow diagram.

**Figure 2:**
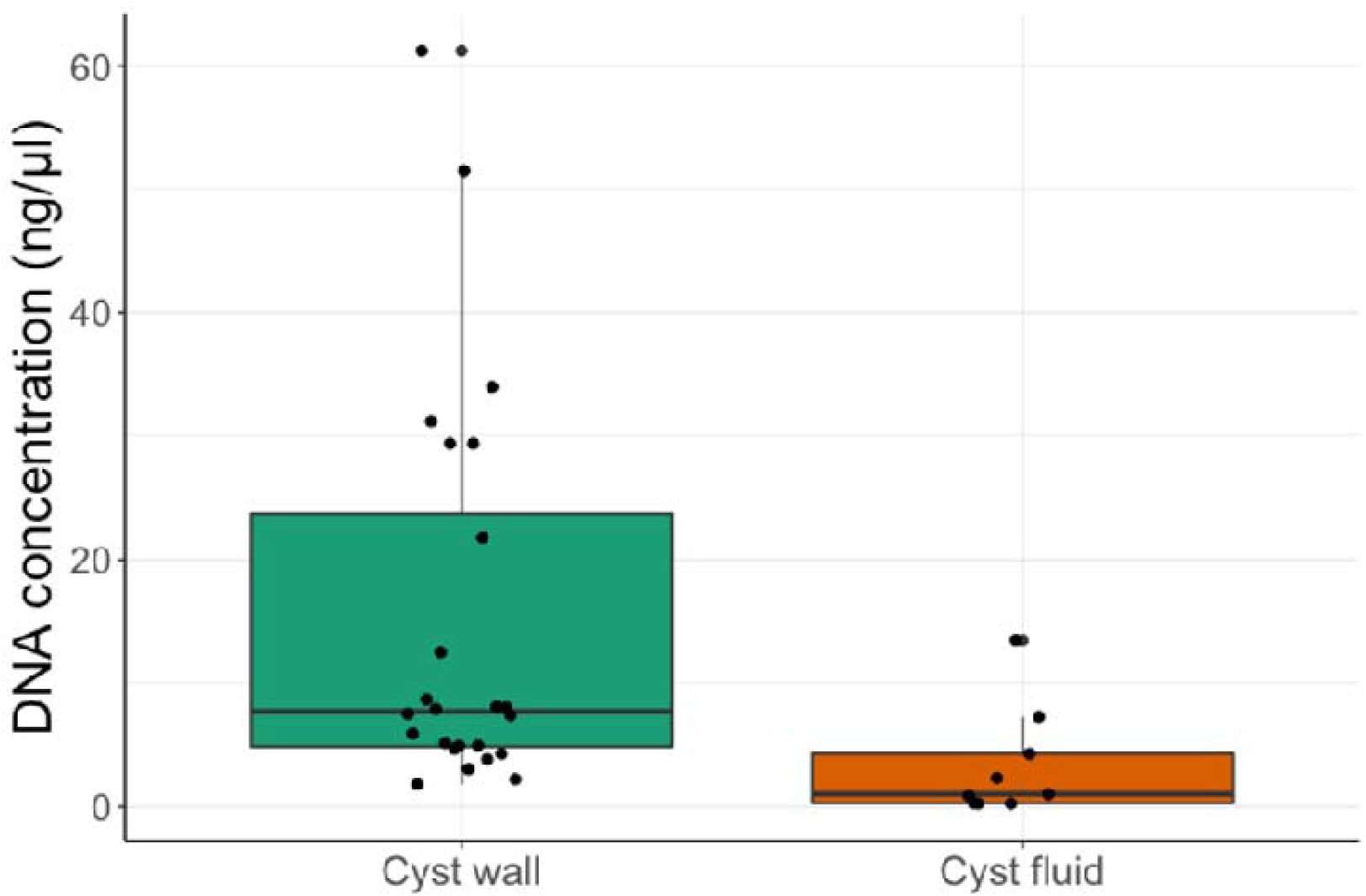
Comparison of DNA concentration (ng/µl) obtained from cyst wall and cyst fluid samples, p=0.0028 Mann-Whitney U test.

### Next-generation sequencing of cyst wall DNA

Mutations in KRAS or GNAS were found in all 7 IPMN samples, and in 2/4 samples graded as probable mucinous cyst. The remaining 2 probable mucinous cysts displayed mutations in TP53. Of the 5 samples with an unknown diagnosis based on histology, 2 carried mutations in KRAS or GNAS and another contained a mutation in TP53. Of the 4 benign cysts, one sample (a pseudocyst) contained a KRAS and VHL mutation. Mutations are summarised in Table 2.

**Table 2:** Somatic protein coding mutations with over 1% prevalence in pancreatic cyst samples. PSC=pseudocyst, EC=epithelial cyst, IPMN=intraductal papillary mucinous neoplasm, SCA=serous cystadenoma, NET=neuroendocrine tumour, LEC=lymphoepithelial cyst, ADC=adenocarcinoma. * indicates a stop codon. All mutant allele frequencies are expressed as percentage of sequence reads containing the mutation compared to the hg38 reference genome wild type sequence.

| Patient | Diagnosis | KRAS | GNAS | TP53 | VHL | NRAS | CTNNB1 | PTEN | AKT1 |
| --- | --- | --- | --- | --- | --- | --- | --- | --- | --- |
| 1 | ADC | p.G12R<br>(10%) | - | - | - | - | - | - | - |
| 2 | NET | - | - | - | - | - | - | - | - |
| 3 | NET | - | - | - | - | - | - | - | - |
| 4 | NET | - | - | - | - | - | - | - | - |
| 5 | IPMN (high grade) | - | p.A844H<br>(31%) | - | - | - | - | - | - |
| 6 | IPMN (low grade) | p.Q61H<br>(40%) | p.R844C<br>(55%) | - | - | - | - | - | - |
| 7 | IPMN (low grade) | p.G12D<br>(8%) | p.R844H<br>(8%) | - | - | - | - | - | p.T172A<br>(1.3%) |
| 8 | IPMN (low grade) | - | p.R844C<br>(3%) | - | - | - | - | - | - |
| 9 | IPMN (low grade) | p.G12D<br>(20%) | - | - | - | - | - | - | - |
| 10 | IPMN (low grade) | p.G12V<br>(6%) | - | - | - | - | - | - | - |
| 11 | IPMN (low grade) | p.G12A<br>(5%) | p.R844H<br>(10%) | - | - | - | p.T41I<br>(6.6%) | - | - |
| 12 | Probable mucinous cyst | - | p.R844H<br>(35%) | - | - | - | - | - | - |
| 13 | Probable mucinous cyst | p.Q61H<br>(2%) | - | - | - | - | - | - | - |
| 14 | Probable mucinous cyst | - | - | p.P85A<br>(2.9%) | - | - | - | - | - |
| 15 | Probable mucinous cyst | - | - | p.V217A<br>(2.5%) | - | - | - | - | - |
| 16 | PSC | - | p.A866G<br>(1.5%) | - | p.L85P<br>(1.5%) | - | - | - | - |
| 17 | EC | - | - | - | - | - | - | - | - |
| 18 | LEC | - | - | - | - | - | - | - | - |
| 19 | SCA | - | - | - | - | - | - | p.Y350H<br>(7%) | - |
| 20 | Unknown | - | - | - | p.R161*<br>(6%) | - | - | - | - |
| 21 | Unknown | - | p.R844H<br>(2.2%) | - | - | - | - | - | - |
| 22 | Unknown | - | - | p.V217A<br>(2.6%) | p.K171*<br>(14.1%) | - | - | - | - |
| 23 | Unknown | p.G12V<br>(4%) | - | - | - | - | - | - | - |
| 24 | Unknown | p.Q61H<br>(11%) | - | - | - | p.Q31G<br>(4.1%) | - | - | - |

### Confirmatory NGS in cyst fluid samples

Confirmatory sequencing was carried out in DNA extracted from cyst fluid on hits in the top 4 genes linked to pancreatic cancer or IPMNs (KRAS, GNAS, TP53, VHL). Variants were confirmed in 4/11 of hits found in cyst wall DNA. A further 2 variants found in the cyst wall DNA were detected in the cyst fluid but failed to pass the Mutect2 quality filter. No additional variants were detected in cyst fluid DNA that were not detected in the corresponding cyst wall DNA.

### Evaluation of the diagnostic accuracy of NGS

The sensitivity, specificity and diagnostic accuracy of EUS + fluid cytology for identifying high-risk cysts were 40%, 100% and 52%, respectively (Table 2). The sensitivity, specificity and diagnostic accuracy of cytology was 12.5%, 100% and 30%, respectively. It was 100%, 25% and 82% for CT/MRI, respectively. The sensitivity, specificity and diagnostic accuracy of standard of care for diagnosing a cyst that needs surveillance or treatment was 66.6%, 50% and 63.1%, respectively. NGS of the cyst wall sample performed better than standard of care with sensitivity of 93.3%, specificity 50%, and diagnostic accuracy of 84.2%. Combining standard of care with NGS improved the above to 100%, 50% and 89.2%, respectively (Table 4).

**Table 3:** Confirmation of GNAS, KRAS, TP53 and VHL variants detected in cyst wall DNA using DNA extracted from cyst fluid. ^†^indicates variant that is present in NGS data but falls below Mutect2 quality filters.

| Patient | Cyst wall variant (MAF) | Present in cyst fluid (MAF) |
| --- | --- | --- |
| 7 | KRAS G12D (8%) | KRAS G12D (45%) |
| 7 | GNAS R844H (8%) | GNAS R844H (39%) |
| 8 | GNAS R844C (3%) | GNAS R844C (7%) |
| 9 | KRAS G12D (20%) | KRAS G12D (40%) |
| 12 | GNAS R844H (35%) | NA<sup>†</sup> |
| 13 | KRAS Q61H (2%) | NA |
| 16 | GNAS A866G (1.5%) | NA |
| 16 | VHL L85P (1.5%) | NA |
| 20 | VHL R161* (6%) | NA |
| 22 | TP53 V217A (2.6%) | NA |
| 22 | VHL K171* (14%) | NA<sup>†</sup> |

**Table 4:** Sensitivity, specificity and diagnostic accuracy of different investigations.

| Investigation | Sensitivity (%) | Specificity (%) | Diagnostic accuracy (%) |
| --- | --- | --- | --- |
| Fluid CEA+ Amylase | 12.5 | 100 | 30 |
| Cytology | 23.08 | 100 | 33.3 |
| EUS+ Cytology | 40 | 100 | 52 |
| NGS | 93.3 | 50 | 84.2 |
| Standard of care (SOC) | 66.6 | 50 | 63.1 |
| NGS+ SOC | 100 | 50 | 89.4 |

## Discussion

This study demonstrates that the yield of extracted DNA is significantly higher from cyst wall tissue samples compared to cyst fluid. The Moray micro-forceps yielded sufficient cyst wall tissue for histological analysis in 62% (15/24) of patients. In contrast, DNA adequate to perform NGS was obtained from all cyst wall samples. The diagnostic accuracy for current standard of care is 63% but the addition of molecular analysis of cyst wall samples improved the diagnostic accuracy to 89%. The cyst wall histology correlated well with the surgical specimens; identified high risk cysts (4/24) that needed surgical intervention and 4/24 had benign cysts who did not need surveillance. However, successful diagnostic histology was obtained only in two-thirds of patients. Molecular analysis of the cyst wall may be an important adjunct in improving the diagnostic accuracy of cysts in the rest of the patients. Hence, histology in conjunction with NGS may help to accurately diagnose high risk pancreatic cysts that need urgent intervention, mucinous cysts that need surveillance and low risk non-mucinous cysts who can be reassured and discharged. Large multi-centre studies with longer follow-up are needed to confirm or refute the above hypothesis.

The low DNA yield obtained from cyst fluid samples is likely due to the highly variable number of cells shed from the cyst. It may also be dependent on the amount of fluid aspirated from the cyst. Obtaining cyst wall tissue with ‘through the needle’ Moray micro-forceps is a feasible method for increasing the DNA yield from pancreatic cysts. It requires minimal training and can be carried out by endosonographers who are trained to do FNA of cysts.

Up to 15% of pancreatic cancers are secondary to mucinous pancreatic cysts [25]. As pancreatic cancers are aggressive and associated with poor survival; the current standard of care is to assess the cyst and survey them with either 6 month or annual CT/MRI scans until the patients become unfit for pancreatic resection [11]. This is associated with increased cost burden to the health care system. It may cause significant anxiety to patients not knowing the type of cysts they have. Molecular analysis of the cyst wall samples with histology may aid to alleviate the above problem.

In this study, the follow up period was relatively short for pancreatic cysts. They were followed up for a median of 491 days. However, during the follow up period there was no overt progression of the cyst lesions. The patients who died had pancreatic adenocarcinoma IPMN with low and high-grade dysplasia. Except for one case of pancreatitis there were no other serious adverse events. This is in line with other studies where the quoted pancreatitis risk following cyst wall biopsy is 3-7%[26]. This is slightly higher than the risk of pancreatitis (2.6%) following FNA of pancreatic cyst[27], although most recent multicentre study using flexible needles reported serious adverse event rate of 1.2% (0.2%-3.5%) [28].

The AmpliSeq panel utilised for this study targets multiple hotspots of 50 genes, these have been narrowed down to genes linked to pancreatic cysts and cancer previously using the COSMIC database [24], and the remainder are included in supplementary data. Mutations in KRAS and GNAS have been proposed as a potential diagnostic test for IPMN but have also been shown to lack the specificity required to detect mucinous cystic neoplasms [29]. Similar to previous studies using cyst fluid [12, 14, 30], we identified KRAS and/or GNAS mutations in 100% of IPMN cases. As expected, all detected KRAS mutations were at codons 12 and 61, the most frequently linked to pancreatic cancer according to the COSMIC database [24]. All but one of the detected GNAS mutations were found at codon 844, which has also been linked to pancreatic cancer and IPMN [31]. A single pseudocyst sample carried a mutation at codon 866, which to our knowledge has not been linked to any cancers. Similar to work in cyst fluid, of the four cysts diagnosed as “probable mucinous cyst” by pathologist, 2 were not found to contain any KRAS or GNAS mutations. TP53 mutations were observed in 3 samples (1 at codon 85 and 2 at codon 217), neither of these sites are associated with pancreatic cancer in COSMIC, but somatic mutations in TP53 at any point have been shown to aid in prediction of outcome of pancreatic adenocarcinoma [32].

We endeavoured to test matched cyst fluid samples from patients where samples could be obtained and sufficient DNA could be extracted. No additional mutations were detected in cyst fluid that were not present in cyst wall, however confirmatory work was limited to amplicons where mutations were already detected, and a full screen with the AmpliSeq panel would be required to confirm this definitively. Interestingly, in two cases inspection of the raw variant call data output from Mutect2 revealed mutations were detected in cyst fluid, before being removed by the default quality filter. To our knowledge this is the first study directly comparing cyst wall and cyst fluid DNA samples from the same patient, and suggests that cyst wall biopsy can provide an increased level of detection of somatic mutations compared to cyst fluid. Additionally, variant allele frequencies were significantly different between cyst wall and cyst fluid. There are several potential explanations for this. Cysts with a greater solid component will shed more DNA into the cyst fluid, this could affect variant allele frequencies simply through differences in the amount of DNA sampled. This bias is avoided by sampling the tissue directly [33]. However, extensive genetic heterogeneity within IPMNs has been demonstrated using single-cell sequencing, meaning results could still be biased depending on the part of the cyst that was sampled [34].

The limitations of the study are recruitment from a single centre, a small sample size and a relatively short follow up period. The authors are aware that this may lead to selection bias and an increased intervention effect, therefore the results cannot be generalised to a wider population. The authors tried to minimise these effects by blinding the histopathologist, radiologist and bioinformatician to patient information. In addition, Moray micro-forceps can only be passed through 19-gauge needle, which may not be feasible in a proportion of cases depending on the site and size of the pancreatic cyst. A recent study demonstrated that use of flexible needle increased the success of EUS-FNA to 89% compared to 75% with the standard needle that we used in this study [28]. To our knowledge, one other study has assessed the utility of NGS of cyst wall biopsies in diagnosis, using formalin-fixed paraffin embedded (FFPE) tissue. Although DNA concentrations were not reported, this study demonstrated that almost 20% of cyst wall specimens recovered from FFPE were unsuitable for NGS, whereas our use of fresh frozen tissue resulted in no excluded samples and a greater average sequencing depth [35].

In conclusion, this study demonstrates that it is feasible to obtain cyst wall samples for histological analysis and NGS. The quantity of DNA obtained from tissue was significantly greater than the cyst fluid samples. Larger studies are needed to assess if NGS and cyst wall histology will aid in accurately characterising cysts as high-risk and requiring intervention, those requiring surveillance, and low risk where patients can be reassured and discharged.

## Data Availability

Anonymised deep sequence data is available from the NCBI Sequence Read Archive under submission SUB11952288.

## Additional information

## Acknowledgements

The authors acknowledge the support of the NIHR Nottingham Biomedical Research Centre (grant BRC-1215-20003). The views expressed are those of the authors and not necessarily those of the NHS, NIHR, or the Department of Health and Social Care.

The authors are grateful to Dr Nadine Holmes and the DeepSeq facility, School of Life Sciences, University of Nottingham for carrying out the NGS, and to Melanie Lingaya, Davor Kresnik, Samatha Warburton, Sian Parkes, Violeta Anuskievic and Rachelle Boxhall for support with sample storage, processing, and training nurses on Moray forceps.

## Authors’ contributions

SA obtained funding, designed the study, acquired and interpreted the data, drafted and revised the manuscript and provided supervision. AB acquired and interpreted the data and drafted the manuscript. JIG obtained funding, designed the study, interpreted the data, revised the manuscript and provided supervision. PK acquired and interpreted the data and revised the manuscript. ADA acquired and interpreted the data and revised the manuscript. MWJ interpreted the data and revised the manuscript. CC acquired and interpreted the data and revised the manuscript. GPA obtained funding, provided supervision and revised the manuscript. SVV obtained funding, designed the study, acquired and interpreted the data, drafted and revised the manuscript and provided supervision.

## Ethics approval and consent to participate

Ethical approval was obtained from the Nottingham Health Science Biobank access committee (REC reference: 15/NW/0685, approval number: ACP000282). All work was carried out in accordance with the Declaration of Helsinki.

## Supplementary data

**Supplementary table 1:** Illumina AmpliSeq Cancer Hotspot Panel V2 primer sequences. Primers used for confirmatory sequencing in cyst fluid are in bold. Available as attached Excel file suppl_table_1.xlsx

